# The Association of Scope of Practice Expansion with Changes in Optometry Workforce Density

**DOI:** 10.64898/2026.08.12.26360334

**Authors:** Dallen Calder, Taylor Johnson, Cameron Carpenter, Charissa Tan, Olaoluwa Omotowa, Chaorong Wu, Phillip Singer, Kathrine Hu, Brian C. Stagg

## Abstract

**Importance:** Access to eye care is increasingly constrained by declining ophthalmologist workforce density, particularly in rural areas, while optometrist workforce is projected to exceed demand. Numerous state legislatures have expanded optometrist scope of practice (SOP), but the workforce effects of these policies have not been evaluated.

**Objective:** To evaluate changes in optometrist and ophthalmologist workforce density following state-level expansion of optometrist scope of practice.

**Design:** This retrospective ecological study analyzed workforce density across 50 states and the District of Columbia. Between 2008–2019, nine states expanded SOPs for optometrists. We used interrupted time-series regression to estimate associations between these policy changes and provider density from 2010-2021, adjusting for covariates.

**Setting:** Population-based analysis of all 50 US states and the District of Columbia, 2010–2021.

**Participants:** State-level workforce data were derived from Bureau of Labor Statistics and US Census data (optometrists) and the American Medical Association Physician Masterfile (ophthalmologists). Socioeconomic covariates were obtained from the American Community Survey.

**Exposure:** State-level legislative expansion of optometrist SOPs to allow injections beyond anti-anaphylaxis treatment, lesion removal, and/or laser procedures. SOP changes were identified through systematic review of legislative and regulatory records.

**Main Outcomes and Measures:** Annual change in optometrist and ophthalmologist workforce density (providers per 100,000 population) following SOP expansion, adjusted for age, income, rates of vision difficulty, diabetes, poverty, and uninsured status.

**Results:** Nine states met inclusion criteria for SOP expansion between 2008 and 2019. Analysis of national data showed that among all 50 US states and the District of Columbia, SOP expansion was associated with a non-significant change in optometrist density (−0.65 per 100,000; 95% CI, −1.87 to 0.57) and ophthalmologist density (+0.09; 95% CI, −0.10 to 0.28). Results were consistent in the 9-state subgroup (optometrists: −0.60, 95% CI −2.90 to 1.70; ophthalmologists: +0.02, 95% CI −0.13 to 0.18).

**Conclusions and Relevance:** Our population-level data suggest that, in the US, state legislation expanding optometrist scope of practice has not been associated with increased workforce density. As numerous state legislatures continue to consider such policies, these findings can inform efforts to balance access to eye care with patient safety.

**Key Points:** *Question:* Does expanding optometrist scope of practice (SOP) increase optometrist workforce density in adopting states?

*Findings:* In this retrospective ecological interrupted time-series study of all 50 US states and the District of Columbia (2010–2021), SOP expansion in 9 states was not associated with a significant change in optometrist or ophthalmologist workforce density.

*Meaning:* Expanding optometrist SOP was not associated with increased workforce density in states that adopted such policies, suggesting that SOP expansion alone may be insufficient to address geographic shortages in eye care access. These findings should be considered by state legislators weighing scope of practice expansion as a workforce solution.

## Introduction

Access to appropriate eye care in the United States, especially in rural areas, is limited by declining ophthalmologist workforce density.^1^ Projections estimate continued shortages as the ophthalmologist workforce declines and the population grows.^2^ In contrast, the density of optometrists has increased and is projected to continue to do so, with supply projected to exceed demand.^1,3,4^

In response, state legislatures across the US are considering laws to allow optometrists to conduct laser and scalpel eye surgery.^5,6^ Proponents of these policies argue that expanded scope of practice (SOP) may incentivize optometrists to relocate to adopting states by offering expanded care roles, opportunities to acquire new skills, and potential for additional billed services.^7,8^ The effect of changing SOPs as a strategy to influence optometry and ophthalmology workforce distribution has not been evaluated. Here we examine the association between state-level optometry SOP expansion and subsequent changes in optometry and ophthalmology workforce density in the US from 2010 to 2021 using publicly available population health databases.

## Methods

We calculated the number of optometrists per state from 2010 to 2021 using data from the Bureau of Labor Statistics (BLS) and the US Census Bureau. Ophthalmologists per capita were calculated similarly using data from the American Medical Association (AMA) Physician Masterfile. State-level SOP changes were identified through a systematic review of legislative records across 50 states and the District of Columbia. An SOP change was defined as an expansion of optometrists’ scope of practice to include injections beyond anti-anaphylaxis treatment, lesion removal, and/or laser procedures. Although implementation of expanded clinical practice varies across states, prior literature suggests that workforce effects tend to emerge gradually rather than immediately, often over a period of one to two years or more.^9,10^ This led us to evaluate workforce effects with a two-year window lag. Thus, our policy review was limited to SOP changes enacted through 2019.

Using interrupted time-series regression with state fixed effects and robust standard errors, we estimated associations between SOP expansion and workforce density, adjusting for temporal autocorrelation and socioeconomic covariates (age, income, vision difficulty, diabetes, poverty, and uninsured status) from the American Community Survey. For visualization, time was expressed relative to SOP change; states without expansion were anchored to 2021 to serve as a comparator group. Analysis was performed in R version 4.5.2.

## Results

Nine states met the study criteria for optometrist scope-of-practice expansion between 2008 and 2019. Alaska (2008), Arkansas (2010), Kentucky (2011), Louisiana (2014), and Virginia (2018) expanded scope of practice to include laser procedures; Georgia (2018) and West Virginia (2010) authorized injections; and Iowa (2009) and Tennessee (2014) authorized lesion removal.

State expanded SOPs were associated with minimal changes in optometrist workforce density. A scatter plot of optometric workforce density over time relative to policy change showed no appreciable change in trend (Figure 1.1). The mean number of optometrists per 100,000 people remained essentially unchanged with 11.5 (sd=3.9) before and 11.3 (sd=4.0) after an SOP expansion (Supplemental Table 1). In contrast, ophthalmologist density decreased after an optometry SOP expansion, declining from 5.8 (sd=2.1) to 4.7 (sd=1.1) per 100,000 population, as illustrated in Figure 1.2.

**Figure 1.1/1.2.**
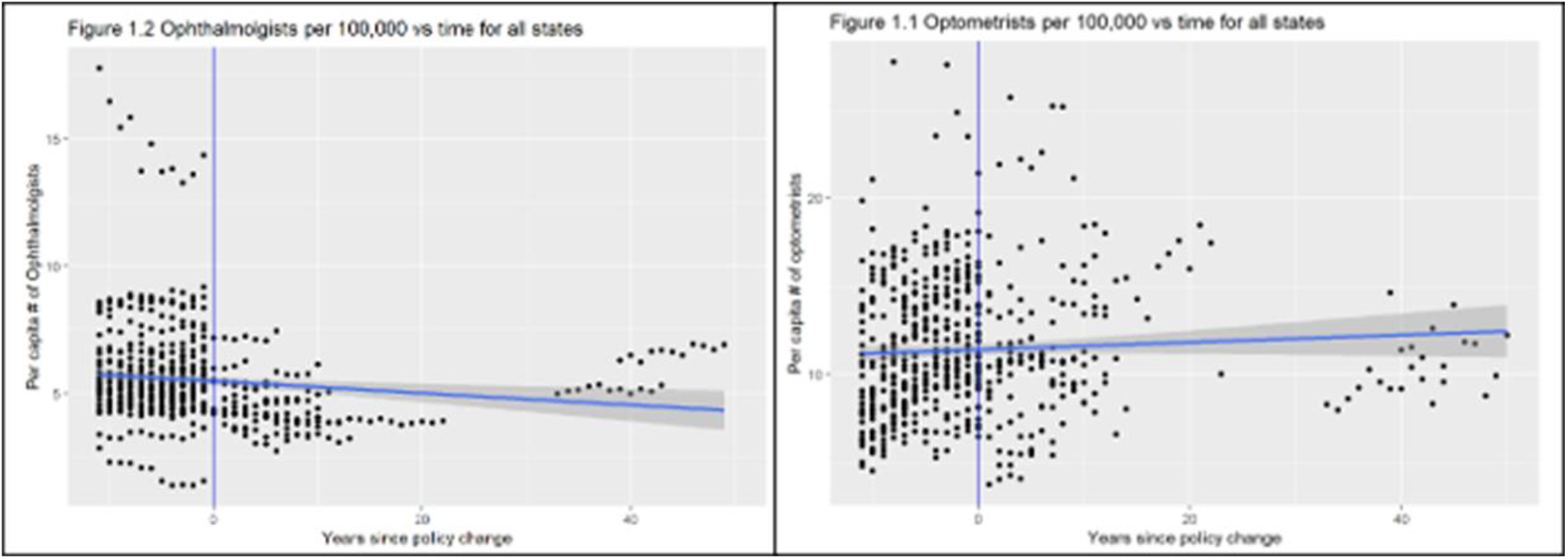
Scatter plots of optometrist (1.1) and ophthalmologist (1.2) workforce density over time relative to policy change, all states. States without an SOP change are anchored to year zero at 2021 and contribute pre-intervention observations only.

**Figure 2.1/2.2.**
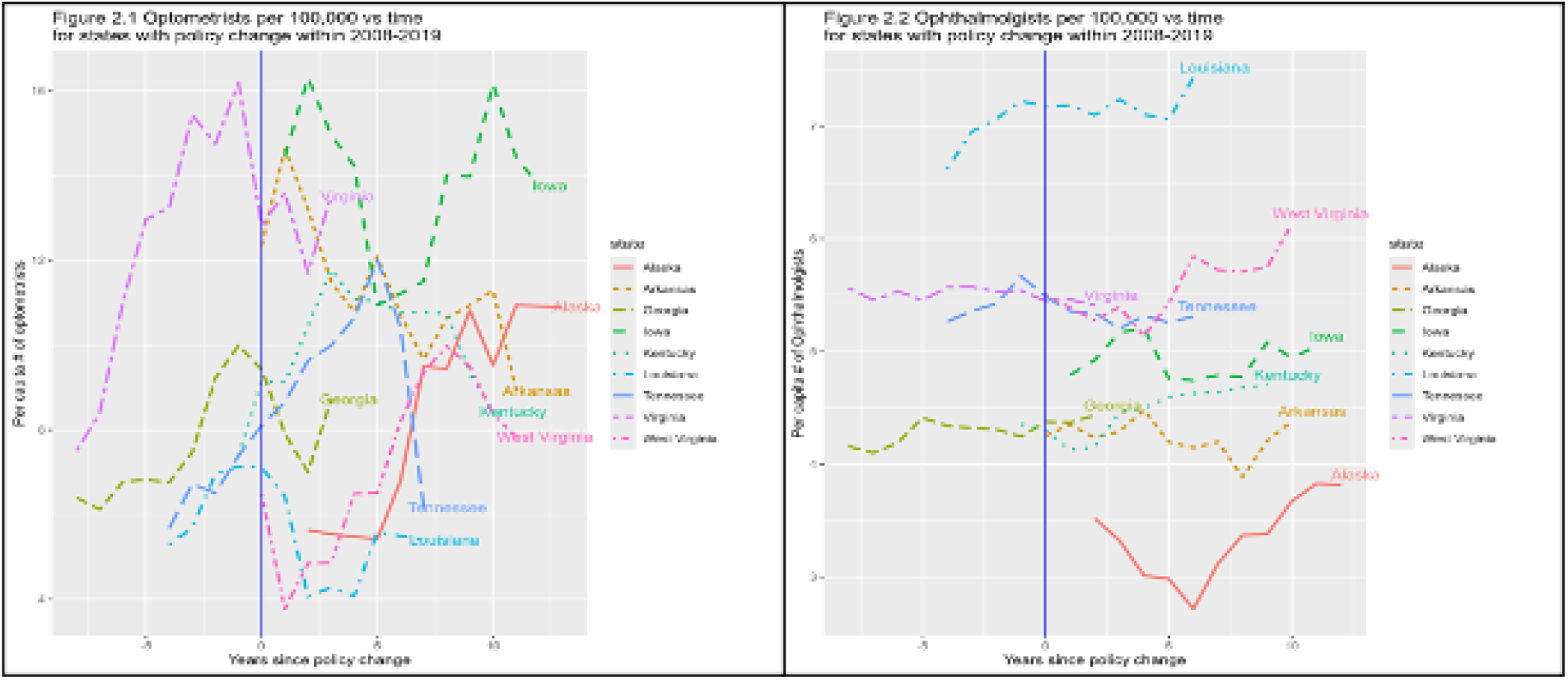
Spaghetti plots of optometrist (2.1) and ophthalmologist (2.2) workforce density relative to policy change for states with SOP expansion within 2008–2019 (n=9). The vertical line denotes the year of policy change.

Figure 2.1/2.2 displays spaghetti plots of workforce density relative to policy change broken down by state for the subgroup (n=9) of states that enacted a policy change between 2008-2019 (shifting forward the time period of interest by two years to give lag time for full potential effects to reveal themselves). Mean optometrist density in this subgroup increased from 8.9 (2.9) per 100,000 before expansion to 11.5 (4.6) after, while ophthalmologists decreased from 5.4 (0.9) to 4.7 (1.0) per 100,000 (Supplemental Table 1).

When controlling for socio-economic and population health characteristics, modeling of both nationwide data and the 9-state subgroup showed that SOP expansion was associated with a non-significant decrease in optometrist density of 0.65 (CI -1.87 – 0.57) and 0.60 (CI -2.90 –1.70) per 100,000, respectively (see Table 1; Supplemental Table 3). The corresponding analysis of ophthalmology data showed another insignificant change, an increase of 0.09 (CI -0.10 – 0.28) in the nationwide group and 0.02 (CI -0.13 – 0.18) in the subgroup.

**Table 1.** Model results for ALL states.

| Predictor | Optometrists per 100,000 |  |  | Ophthalmologists per 100,000 |  |  |
| --- | --- | --- | --- | --- | --- | --- |
|  | Estimate | 95% CI | p | Estimate | 95% CI | p |
| (Intercept) | 15.29 | -0.24 – 30.82 | 0.054 | 2.29 | -0.66 – 5.25 | 0.128 |
| time | 0.07 | -0.01 – 0.14 | 0.100 | -0.02 | -0.04 – 0.00 | 0.115 |
| intervention [1] | -0.65 | -1.87 – 0.57 | 0.296 | 0.09 | -0.10 – 0.28 | 0.334 |
| VisionD | -14.83 | -109.02 – 79.36 | 0.757 | -21.66 | -34.16 – -9.16 | **0.001\*\*** |
| diabetes | -0.21 | -0.54 – 0.11 | 0.198 | 0.01 | -0.04 – 0.06 | 0.733 |
| income | 0.00 | -0.00 – 0.00 | 0.900 | -0.00 | -0.00 – -0.00 | **<0.001\*\*\*** |
| age | 0.09 | -0.29 – 0.46 | 0.656 | 0.12 | 0.05 – 0.19 | **0.001\*\*** |
| uninsured | -14.53 | -26.41 – -2.64 | **0.017**<br>* | -2.22 | -3.85 – -0.60 | **0.007\*\*** |
| povertyrate | -0.23 | -0.46 – 0.00 | 0.055 | 0.00 | -0.04 – 0.04 | 0.980 |
| **Random Effects** |  |  |  |  |  |  |
| $\sigma^2$ | 5.50 | | | 0.08 | | |
| $\tau_{00}$ (state) | 7.82 | | | 3.89 | | |
| ICC | 0.59 |  |  | 0.98 |  |  |
| N (states) | 51 |  |  | 51 |  |  |
| Observations | 537 |  |  | 508 |  |  |
| Marginal R<sup>2</sup> /<br>Conditional R<sup>2</sup> | 0.170 | / 0.657 |  | 0.031 | / 0.981 |  |

\* $p < 0.05$ \*\* $p < 0.01$ \*\*\* $p < 0.001$

## Discussion

We find little evidence that expanding optometry SOPs increases provider density. These results were consistent across unadjusted and adjusted models. No statistically significant association between provider density and SOP expansion was found while controlling for demographic, need, and economic variables. The anticipated workforce response to SOP expansion did not materialize.

A possible explanation for these findings is the predominantly rural nature of many states that expanded optometrist SOPs. Rural areas have historically experienced slower growth in eye care provider density than urban areas, regardless of policy. ^1^ Among the nine states in our analysis, five rank among the 15 most rural states in the United States by population. ^11^ Although we did not directly evaluate this mechanism, persistent challenges in recruiting providers to rural areas may limit the workforce effects of scope-of-practice expansion.

In 2025, 16 state legislatures introduced legislation to expand optometrist scope of practice.^12^ Few studies have evaluated the impact of SOP expansion on patient care and safety. One of the only studies in this area found that patients undergoing laser trabeculoplasty (LTP) were much more likely to require additional LTPs.^13^ While more research in this area is needed, our findings can inform legislators in their efforts to balance access to eye care with patient safety.^14,15^

## Limitations

Several limitations should be considered. First, different types of SOP expansions (lasers, injections, and lesion removal) were treated as equivalent, though their clinical and recruitment impacts may differ. Second, the analysis focused only on a state’s initial expansion. Third, the reliance on administrative and legislative databases introduces inherent variability. Fourth, while a 2-year lag is supported by literature^, 9, 10^ a longer follow-up might be required to observe significant workforce shifts. Finally, the small sample size of nine intervention states limits the statistical power of the subgroup analysis

## Conclusion

Our population-level analysis suggests that expanding optometry scope of practice has not been associated with increased workforce density in the US. Although national trends in both optometry and ophthalmology were observed over time, recent scope of practice expansions were not linked to meaningful changes in provider density. These findings may help inform legislative discussions regarding strategies to improve access to eye care.

## Data Availability

All data produced in the present study are available upon reasonable request to the authors

## Supplemental Materials

**Supplemental Table 1.** Before and after policy change for all states.

|  | Before Policy Mean (sd) | After Policy Mean (sd) |
| --- | --- | --- |
| Total N (%) | 403 (65.8) | 209 (34.2) |
| Optometrists per 100,000 | 11.5 (3.9) | 11.3 (4.0) |
| Ophthalmologists per 100,000 | 5.8 (2.2) | 4.8 (1.0) |

**Supplemental Table 2.** Before and after policy change for states with policy change within 2008-2019.

|  | Before Policy Mean (sd) | After Policy Mean (sd) |
| --- | --- | --- |
| Total N (%) | 33 (34.4) | 63 (65.6) |
| Optometrists per 100,000 | 10.3 (5.4) | 9.8 (3.1) |
| Ophthalmologists per 100,000 | 5.5 (0.9) | 5.1 (1.0) |

**Supplemental Table 3.**
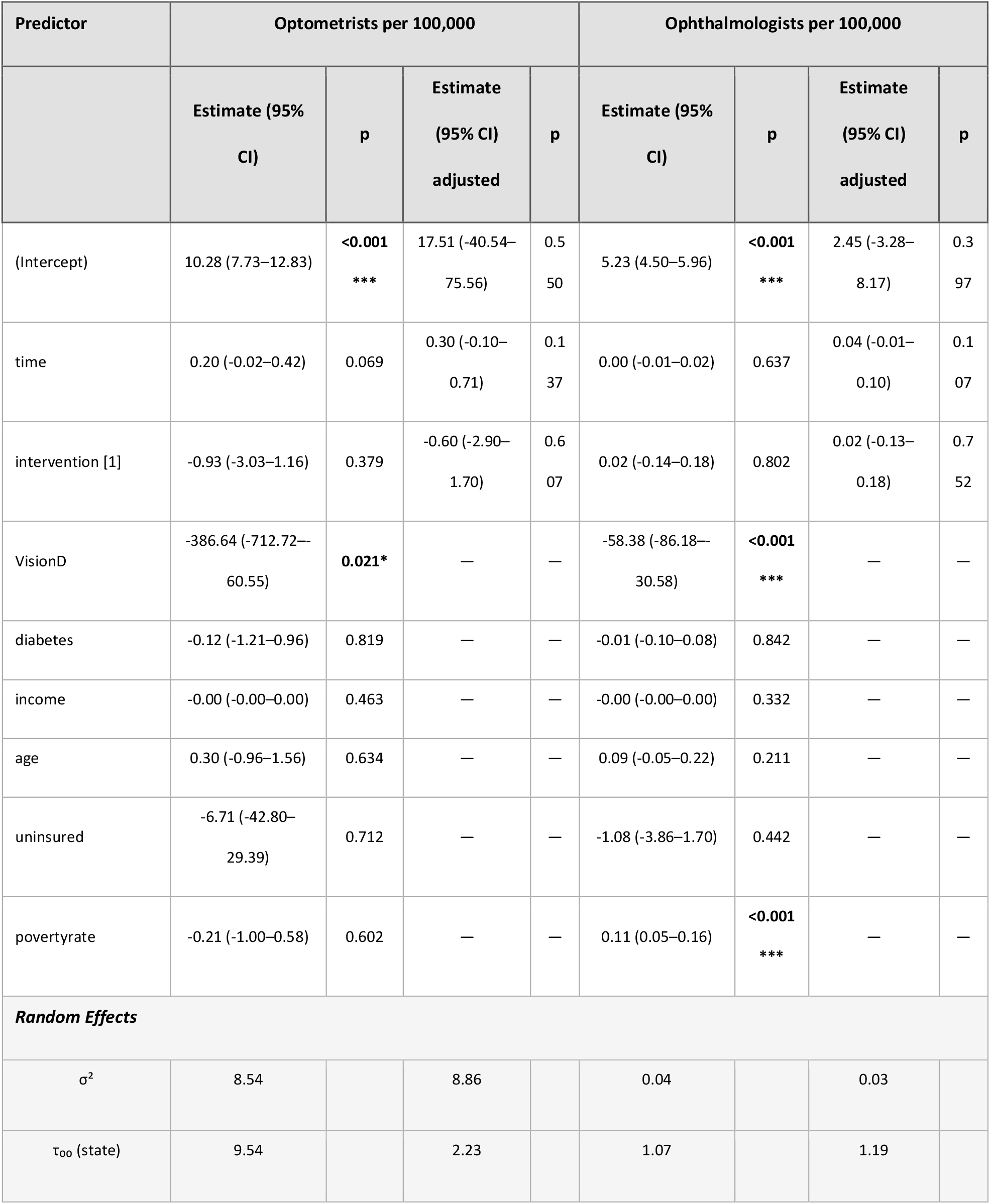
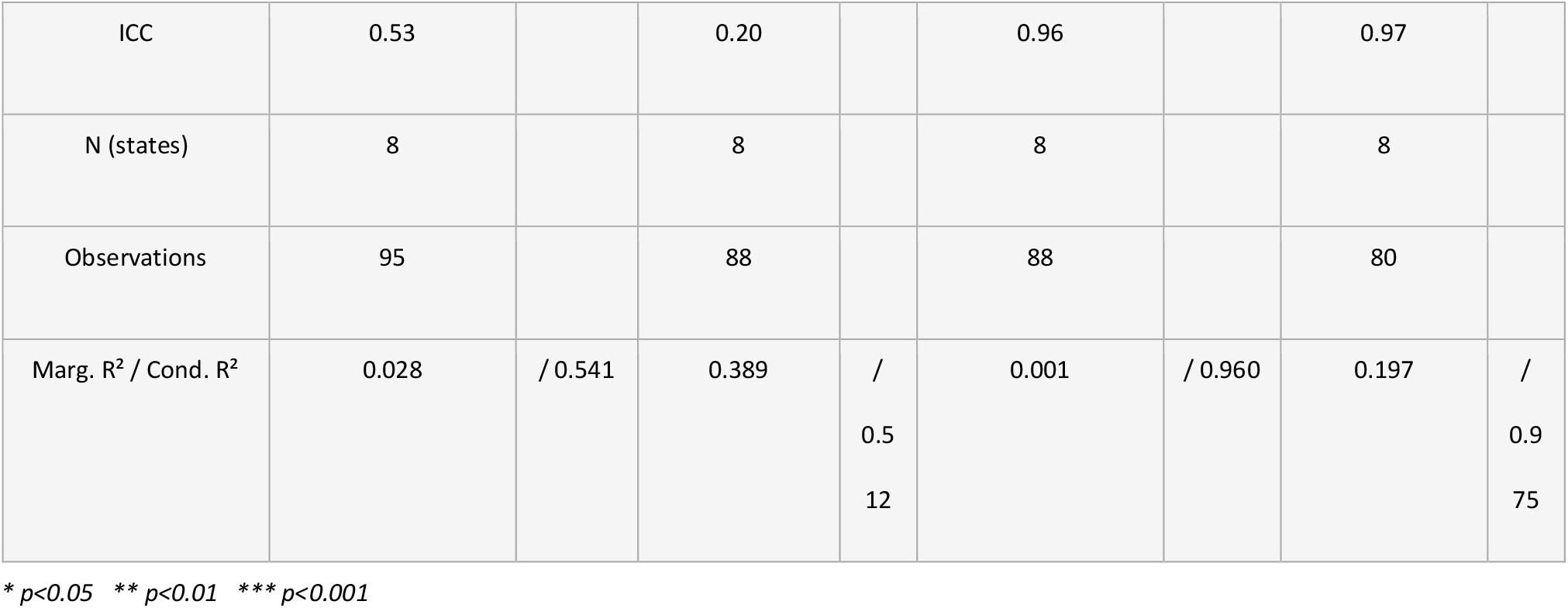
Model results for states with policy change during 2008-2019.

